# High connectivity and human movement limits the impact of travel time on infectious disease transmission

**DOI:** 10.1101/2023.07.26.23293210

**Authors:** Reju Sam John, Joel C. Miller, Renata L. Muylaert, David T. S. Hayman

## Abstract

The speed of spread of severe acute respiratory syndrome coronavirus 2 (SARS-CoV-2) during the coronavirus disease 2019 (COVID-19) pandemic highlights the importance of understanding how infections are transmitted in a highly connected world. Prior to vaccination, changes in human mobility patterns were used as non-pharmaceutical interventions to eliminate or suppress viral transmission. The rapid spread of respiratory viruses, various intervention approaches, and the global dissemination of SARS-CoV-2 underscore the necessity for epidemiological models that incorporate mobility to comprehend the spread of the virus. Here, we introduce a metapopulation susceptible–exposed–infectious–recovered (SEIR) model parameterised with human movement data from 340 cities in China. Our model replicates the early case trajectory in the COVID-19 pandemic. We then use machine learning algorithms to determine which network properties best predict spread between cities and find travel time to be most important, followed by the human movement Weighted Personalised PageRank. However, we show that travel time is most influential locally, after which the high connectivity between cities reduces the impact of travel time between individual cities on transmission speed. Additionally, we demonstrate that only significantly reduced movement substantially impacts infection spread times throughout the network.

## 1. Introduction

Severe acute respiratory syndrome coronavirus 2 (SARS-CoV-2) emerged in late 2019 and spread throughout the world in 2020 [1, 2]. Computational epidemiological modeling has been an important tool to predict the emergence and propagation of the pathogen [3]. The role of travel in infection spread is well recognised and pathogens can now move faster than ever before due to modernisation and globalisation [4]. Hence, travel restrictions are a key, non-pharmaceutical tool for ceasing or slowing the transmission of pathogens between locations [5, 6]. For an effective implementation of such interventions we need a better understanding of the effects and benefits of such restrictions *a priori*. This calls for a metapopulation modeling approach that incorporates population exchange between different locations.

Basic metapopulation models do not identify individuals based on their home locations and are formulated as Markovian processes. However, the primary pattern of human movement is driven by commuting populations. Thus, it is important to track the travellers according to their original locations. Hence we developed a metapopulation susceptible-exposed-infected-recovered (SEIR) compartmental model for commuting individuals in a population, using population and mobility data at the city level.

Researchers have built different epidemiological models to study the spread of SARS-CoV-2 (e.g. [3, 7, 8]). Few models take into account empirical population flows, which are key for understanding transmission dynamics [3, 6, 8–12]. Such models help to accurately depict the actual dynamics of the spread, which are highly influenced by the population flow in a real-life setting [3, 12]. Previous work has applied a global metapopulation disease transmission model with some national subpopulations centered around major transportation hubs, including within China, to model the impact of travel limitations on the national and international spread of SARS-CoV-2 after its emergence in Wuhan, China [12]. These models, however, have focused on SARS-CoV-2 dynamics and spread in the face of a pandemic, but not the overall dynamics of the systems and properties of the networks that facilitate or limit spread. As a result, there is a pressing need for data-driven, quantitative studies that look at fundamental system properties to help plan for future outbreaks for infections with similar epidemiological features.

Here, we created a model for the transmission of a SARS-CoV-2-like infection, using population flow data to effectively describe the infection spread. We used population flow data from all cities in mainland China to their top 100 most connected cities to constrain the parameters in our model. We built the population flow network of China with cities as nodes and the population flow between the cities as the edge weight of the nodes. We first consider models with introductions of 100 infections in Wuhan, using this as the epicentre of the SARS-CoV-2 outbreak, [13], and simulate the transmission behavior and outbreak sizes of SARS-CoV-2-like infections across China, then seed infection in different localities. We investigated which network properties are most important in the spread of infection, and whether there is a relationship between commuter numbers and their travel duration, infectious period, basic reproduction number *R*_0_ (the number of cases generated by a typical index case in a fully susceptible population), and incubation period of the infection. Then, we conducted simulations to understand the impacts of reducing human flow between cities as a non-pharmaceutical intervention to control spread between cities.

## 2. Methods

### (a) Model structure

We developed a metapopulation SEIR model to study the transmission dynamics of an emerging COVID-19 like disease. We assigned home locations to each human host, identified as the level 2 administrative division in China. Let us imagine we have *n* such locations. The people usually live in a particular location and travel occasionally or periodically to other locations (in a temporary basis). Here we differentiate between the residents and visitors currently located at the same location. Therefore we label *S*_*ii*_(*t*), *E*_*ii*_(*t*), *I*_*ii*_(*t*), *R*_*ii*_(*t*), and *N*_*ii*_(*t*) as the number of susceptible, exposed, infected, recovered and total people in location *i* at time *t* who belong to location *i* and *S*_*ij*_ (*t*), *E*_*ij*_ (*t*), *I*_*ij*_ (*t*), and *R*_*ij*_ (*t*) as the number of susceptible, exposed, infected, and recovered people in location *i* at time *t* who belong to location *j*. These visitors are represented by the dashed circles in Figure 1. The visitors interact with residents at a given location and they return to their home location at a fixed rate *π*_*ij*_. If people from location *j* travel to location *i* and stay there for a week before returning to their home location *j*, then the return rate *π*_*ij*_ will be 1*/*7 per day. For simplicity, we assume the same return rate (*π*) for everyone at all locations. The total hosts currently in location *i* that live in location *j* will then be, *N*_*ij*_ (*t*) = *S*_*ij*_ (*t*) + *E*_*ij*_ (*t*) + *I*_*ij*_ (*t*) +*R*_*ij*_ (*t*). The basic *SEIR* model structure which incorporates the movement and vital dynamics (birth and natural death) is shown in Figure 1.

**Figure 1.**
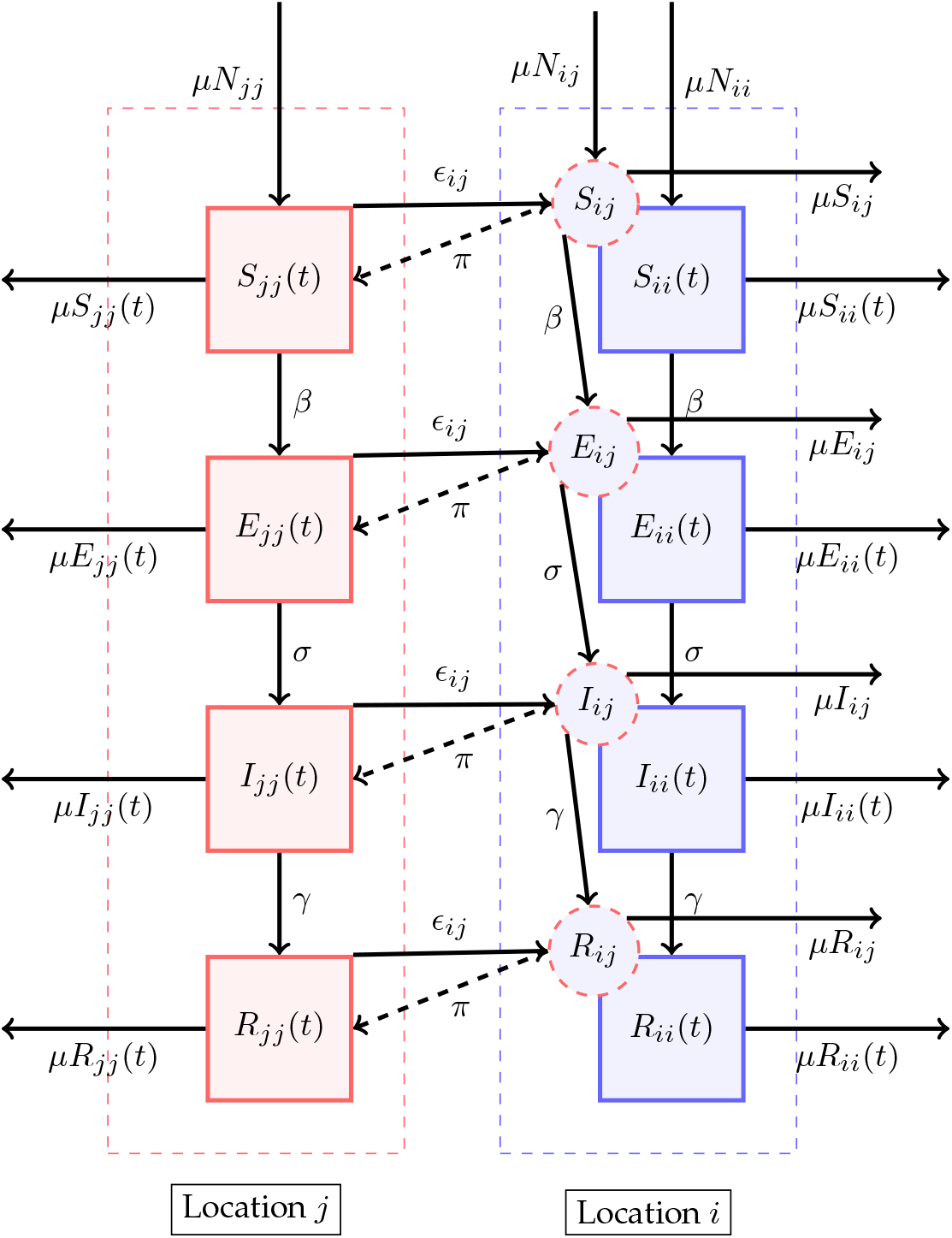
The schematic representation of the *SEIR* metapopulation model. In the compartment diagram, inhabitants of a particular city or location are represented by a particular color. The square boxes represent the set of individuals who are in the Susceptible, Exposed, Infectious, and Recovered class. The downward arrows represent the flow from one compartment to another within a city or population and are annotated with the corresponding flow rates. The arrow pointing to the right between the square boxes represents the flow between the corresponding compartments of another city or population and is annotated with the emigration rate. The dashed circles represent immigrants in a particular city, for example, *j*, who emigrated from another city, for example, *i*. The dashed arrow represents the return of immigrants to the home city and is annotated with the return rate. The outward and inward arrows from and to the square boxes represent death and birth, annotated with the respective death and birth numbers, assuming the same birth and death rates.

Let *ϵ*_*ij*_ represent the rate at which hosts whose home city is *j* travel to *i*, with *ϵ*_*ii*_ = 0 for all *i*. A person from location *j* who is currently at the location *i* will be included in the *N*_*ij*_ population. The number of people who belong to the location *i* remains constant over time, even though the members visit other sites 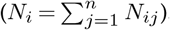. Further, to make this statement true we made another assumption: birth rate, *μ* = death rate, *μ* (so, *μN*_*ii*_(*t*) = *μS*_*ii*_(*t*) + *μE*_*ii*_(*t*) + *μI*_*ii*_(*t*) + *μR*_*ii*_(*t*)). Finally, we assume homogeneous mixing for individuals within each city. With these assumptions the 8*n*^2^ equations describing transmission among the peoples while they are at their home site or while they are travelling are:

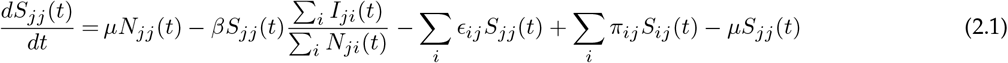

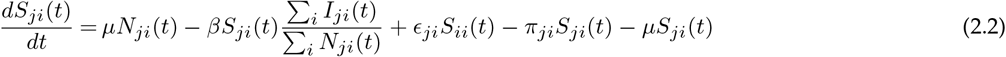

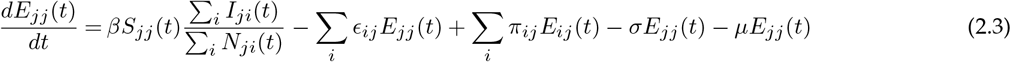

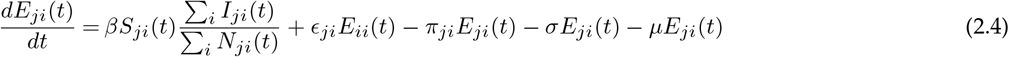

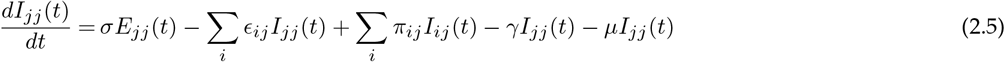

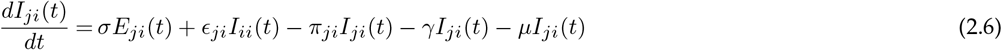

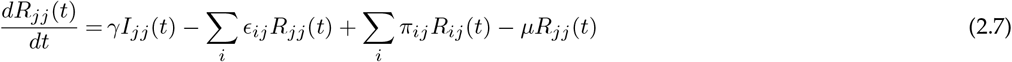

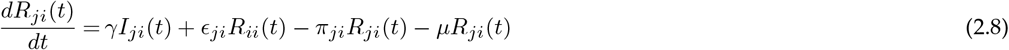

where the parameters are summarised in Table 1.

**Table 1:** Parameters and variables.

|  | Description | Values | Units | Refs. |
| --- | --- | --- | --- | --- |
| $R_0$ | Basic reproduction number | 3 | Dimensionless | [14]* |
| $\mu$ | Birth & death rate | $1/(60 \times 365)$ | Per capita day<sup>-1</sup> | [15] |
| $\sigma$ | Latency rate | $1/5.5$ | Day<sup>-1</sup> | [16, 17] |
| $\gamma$ | Recovery rate | $1/10$ | Day<sup>-1</sup> | [15] |
| $\beta$ | Transmission coefficient | $R_0(\gamma + \mu)$ | Per capita day<sup>-1</sup> | [18] |
| $\epsilon_{ij}$ | Emigration rate | Estimated from Baidu server | Per capita day<sup>-1</sup> | NA |
| $\pi$ | Return rate | $1/5$ | Day<sup>-1</sup> | NA |

\*Liu *et al.* compared 12 published studies and estimated the $R_0$ to range from 1.5 to 6.68. We rounded their mean value of 3.28 to 3.

### (b) Human movement data

We need to estimate the emigration rate, *ϵ*_ij_ for the populations from all the cities in China. For this we scraped the Baidu migration site (http://qianxi.baidu.com/) [19] for a period from January 19, 2021 to January 18, 2022. The Baidu migration data set is created from a mobile phone application that tracks the movements of users. The Baidu migration website provides the 100 most popular immigration sources and emigration destination locations of each prefecture administrative level. The immigration/emigration (*η*) of a city is provided as the percentage of the population that migrated from/immigrated to the corresponding city. The website also displays the inward/outward migration index (hereafter Baidu migration index – *ι*) of all administrative divisions in the mainland China. Hence, the real inward/outward migration can be calculated as,

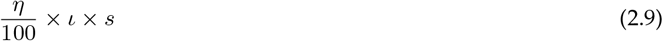

Where the number *s* is the scaling factor that converts the Baidu migration index (*ι*) to the absolute number of travellers. However, the numerical value of this scale is ambiguous. Several authors have calculated different values for each unit of Baidu’s migration index, which is summarised in Table 2. Combining evidence from the sources in Table 2, we chose a scaling factor of 50, 000.

**Table 2:** Various scaling factors for Baidu migration index (*ι*) from different sources.

| Reference | $\iota$ index scale |
| --- | --- |
| Jiang et al. 2021 [20] | 33, 333.00 |
| Tian et al. 2020 [21] | 40, 926.89 |
| Sanche et al. 2020 [22] | 44, 519.62 |
| Yuan et al. 2020 [23] | 56, 137.00 |
| Pengyu et al. 2022 [24] | 71, 121.00 |

### (c) Human flow network

We use the location-based service offered by the Baidu data server, which gathers information based on Global Positioning System (GPS) locations, locations of cell towers, IP addresses, Wi-Fi, and location data from a variety of software and apps on mobile devices. We collected the mobility data by monitoring the features of the HTTPS (Hypertext Transfer Protocol Secure) requests made to the Baidu data server. This provides us the percentage of movement for all cities and their one hundred most connected cities. After analysing the responding JavaScript Object Notation (JSON) file from the server, the outflow and inflow matrix for the cities in China is generated. Then we took the yearly average of this inflow and outflow matrix, and it turned out that one matrix is approximately the transpose of the other which corresponds to almost all travel being round trips. Therefore, we construct the parameter, *ϵ* in the model as: *ϵ* = (Inflow + Outflow^*T*^)/2. Table 3 presents a sample of the yearly average flow matrix (*ϵ*) between 8 example cities.

**Table 3:** Sample of the yearly average humna population flow matrix. Eight cities are shown from 340 in total.

| City | Beijing | Tianjin | Shijiazhuang | Tangshan | Qinhuangdao | Handan | Xingtai | Baoding |
| --- | --- | --- | --- | --- | --- | --- | --- | --- |
| Beijing | 0.00 | 29088.15 | 8023.07 | 8039.97 | 4858.53 | 6970.73 | 3650.78 | 32726.33 |
| Tianjin | 28971.87 | 0.00 | 3160.28 | 18913.49 | 2983.07 | 4529.83 | 1599.47 | 6247.30 |
| Shijiazhuang | 8043.85 | 3152.62 | 0.00 | 2572.41 | 1163.29 | 8026.34 | 16629.05 | 17161.73 |
| Tangshan | 8003.41 | 18897.44 | 2572.60 | 0.00 | 9155.63 | 828.69 | 565.14 | 2128.62 |
| Qinhuangdao | 4835.46 | 2973.81 | 1161.39 | 9150.64 | 0.00 | 271.74 | 214.46 | 948.24 |
| Handan | 6920.69 | 4520.10 | 8029.35 | 828.20 | 272.98 | 0.00 | 12145.52 | 2100.95 |
| Xingtai | 3657.15 | 1592.72 | 16627.32 | 564.59 | 215.30 | 12125.10 | 0.00 | 1919.13 |
| Baoding | 32584.71 | 6236.65 | 17175.34 | 2128.95 | 951.33 | 2101.84 | 1922.95 | 0.00 |

After developing the infectious disease spread model for the country, simulation experiments were carried out to identify the key parameters that can affect epidemic spread in a highly connected country like China. One of the main parameters that affects epidemic spread is the number of initial infected people in a population. For performing a systematic study to identify the key parameters that can affect epidemic spread, we kept a constant 100 initial infected people throughout the experiment.

### (d) Model validation

According to Huang et al., 2020 [25] and Allam 2020 [26], the earliest date of reported COVID-19 cases at Wuhan in Hubei province of China was December 1, 2019, though the earliest infections and cases are not known (see [13]). However, this date is reasonable approximation for our purposes of tracking early infection dynamics. As of February 10, 2020, approximately 71 days after the first case in Wuhan, there had been 262 confirmed cases of COVID-19 in Beijing. [27]. We use these numbers and time frame to help validate the model.

### (e) Predictors of spread

There are important features of networks that might govern epidemic spread. These include factors relating to the flow of people, such as the number of travellers (see above), the travel duration and distance, and specific network (graph) properties. Here, we identify key metrics and test which ones influence the spread in our metapopulation which we now describe below.

#### (i) Travel duration

To understand the effect of travel time, we retrieve and analyze road/street networks of China from the OpenStreetMap (OSM) with OSMnx [28]. A known node in the created China road network is a location of interest on the map, such as a bus stop, house, shop, or train station. The roads that connect these nodes are our edges. They have some useful metadata like distance and the maximum speed allowed on that particular road. We defined the centroid of each prefecture-level city (China administrative level 2) through performing a spatial match between the location reference codes from Baidu and a reference polygon shapefile [29] of China compiled by the United Nations Office for the Coordination of Humanitarian Affairs (OCHA) and the Regional Office for Asia and the Pacific (ROAP). Then we find a known node close to the centroid of each prefecture-level city polygon. This enables us to identify a shortest route between Beijing and all the city centers by connecting all the intermediate connected nodes. One such shortest possible route between Wuhan and Beijing is shown in Figure 2.

**Figure 2.**
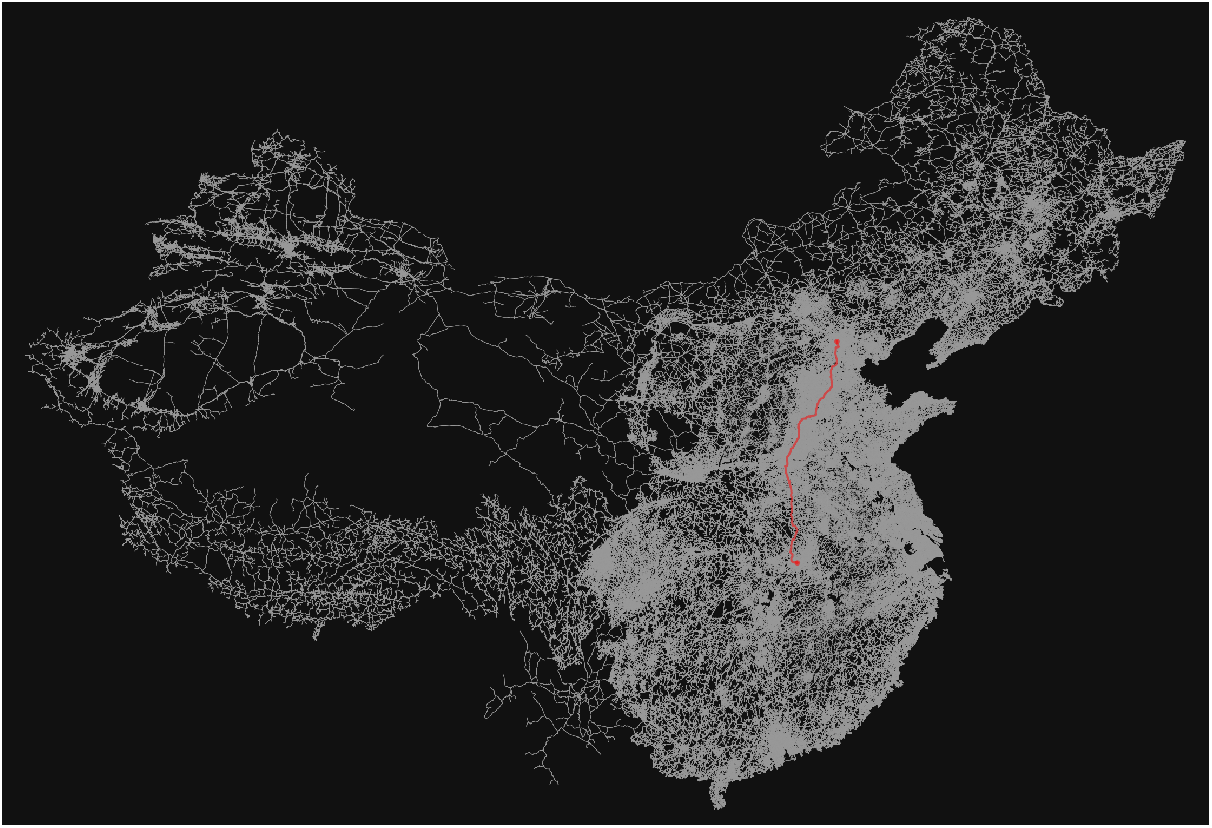
Shortest possible route between Wuhan and Beijing centroids (red) calculated from the China road network (white)

Then we calculated the total distance and travel time between city centers and Beijing by adding the distance between all the nodes between those points. When we compared the drive (motorized-vehicle travel time) time calculated from OSM with Google maps, it appears that OSM is always very (too) fast, even though the distances are comparable. This is due to the fact that the time calculation is done by adding the time to traverse each edge with the assumption that one can travel at the maximum speed limit of that road (edge). For a better understanding of the human flow (immigration and emigration) and infectious disease spread among cities in China, we created another graph of China, where each node is a city and edges between them are weighted by the actual flow of people that we calculated from *ι*.

#### (ii) Network statistics

To compute the network statistics for the China travel network, we created a graph of China, where each city is represented as a node and the flow/outflow matrix between each city is used to create the edges between those nodes, which are weighted with the number of people moving between those cities. There are multiple measures of association between nodes in a graph. These measures include node-level ranking algorithms using link-based centrality metrics, including Google’s PageRank, Degree Centrality, Betweenness Centrality and others. We use eight metrics, which are described below, to explore the role of different network properties on spread in our metapopulation.

**Degree Centrality** is a measure of the connectivity of a city within a network [30]. It is calculated by counting the number of edges a city has and normalizing it by dividing it by the maximum degree in the graph. This measure gives us an understanding of how many cities a particular city is connected to through human mobility and, therefore, how influential it is within the network.

**Eigenvector Centrality** is a measure of the influence of a city within a network [30–32]. It takes into consideration the centrality of the cities pointing to a particular city and assigns a higher eigenvector centrality value to cities that are visited by people from many other central cities. In our analysis, we reversed the directional graph of the China outflow network to get a better understanding of the influence of a city based on the number of cities it is pointing to, within the eigenvector centrality analysis.

**PageRank** is an algorithm that computes a ranking of nodes (here city) in a graph based on the structure of incoming links [33, 34]. It has several possible improvements over other centrality measures, such as eigenvector centrality. In the PageRank algorithm that we followed([35]), every node (city) has an arbitrary amount of centrality at the outset. Hence, even an unlinked node will have a baseline centrality; that is, a city’s existence itself gives it an alpha centrality. Also in PageRank, if we have two cities with the same centrality measure, the one with fewer outflow links will transfer more value to the linked nodes than the other.

**Weighted PageRank** is a modification of the PageRank algorithm that takes into account the weight of edges in a graph, where in our analysis the edge weight is the number of people flowing out between the cities [34, 36]. This modification provides a more accurate picture of the spread of information or people in a network.

**Weighted Personalized PageRank**, also called “Random Walk with Restart” [37–39], is a variant of the Weighted PageRank algorithm for finding nodes in a graph that are most relevant to another node. Here Weighted Personalized PageRank is adapted for both the outflow from Wuhan and the flow to Beijing [40]. This modification provides a more accurate understanding of the gravity of a node in the graph and how fast Beijing can reach 100 infections if Wuhan has an initial 100 infections.

The **HITS** [41, 42] algorithm, also known as “hubs and authorities”, is an alternative method of identifying relevant and popular nodes in a network. It provides two separate measures: Authority score and Hub score. The Hub score is calculated by collecting links from the nodes linked to a particular node and assigning a score based on the number of links received and from which nodes. This measure provides a better understanding of the relevance of a city in the network.

**Betweenness Centrality** is a measure of how often a shortest path between all possible connected nodes passes through a particular node [30, 43–46]. This measure gives us an understanding of the role of a node as a bridge between different parts of the network, facilitating the flow of people from one part of the network to another. A city with high Betweenness Centrality is a critical component of network connectivity, and a decrease in the Betweenness Centrality of a city could significantly impact the flow of people through that city.

### (f) Variable importance

#### (i) Principal component analysis (PCA)

Principal component analysis (PCA) allows us to analyse and visualize multivariate data. We used PCA to allow us to see the relative contribution of the different metapopulation network properties of each node (here city) relating to infection spread and help determine which properties to use in further statistical analyses. PCA was performed using the tool referenced in [47].

#### (ii) Machine learning algorithms

Because of the non-linear relationships between location properties and the time it took for 100 cases to reach Beijing (our response metric; see Results), we used two machine learning approaches to understand which variables are most important in predicting spread. We used gradient boosting regression tree (BRT) and random forest (RF) analyses, both through ensemble approaches. These approaches have different overfitting diagnosis and accuracy properties [48]. We only used the metrics that were not highly correlated (see Results); travel time, Weighted Personalized PageRank (Beijing flow), Weighted Personalized PageRank (Wuhan Outflow), Population, Degree Centrality, PageRank, and Betweenness Centrality.

### (g) Movement and interventions

An important parameter that we hypothesized would alter the epidemic transmission dynamics is the return rate, *π*, of the commuting population. To estimate its effect, we set up an experiment where we varied the return rate from 1*/*1 to 1*/*30.

Lastly, several analyses have looked at the impact of non-pharmaceutical interventions, such as ‘lockdowns’ [5, 8, 49]. In this study, we replicate these interventions to determine the necessary reduction in human flow in a data-driven model. We accomplish this by first reducing the daily population flow from Wuhan and measuring the time it takes to reach 100 cases in Beijing. Then, we reduce the daily population flow from Wuhan and the top five locations linked to Wuhan identified by Weighted Personalized PageRank (Wuhan outflow). Another experiment that can be performed is by reducing the daily population flow from Wuhan and the top five locations identified by the weighted personalized PageRank value for Beijing flow, since it is the second most important metric determining the epidemic spread to Beijing, as one can infer from the BRT and RF analyses (see Results).

## 3. Results

Our simulation of the model with the parameters listed in the Table 1 above gives similar estimates of the epidemic size and timing of the early stages of the COVID-19 outbreak in Beijing, reaching 100 cases in Beijing on day 60 [25–27] (Figure 3a).

**Figure 3.**
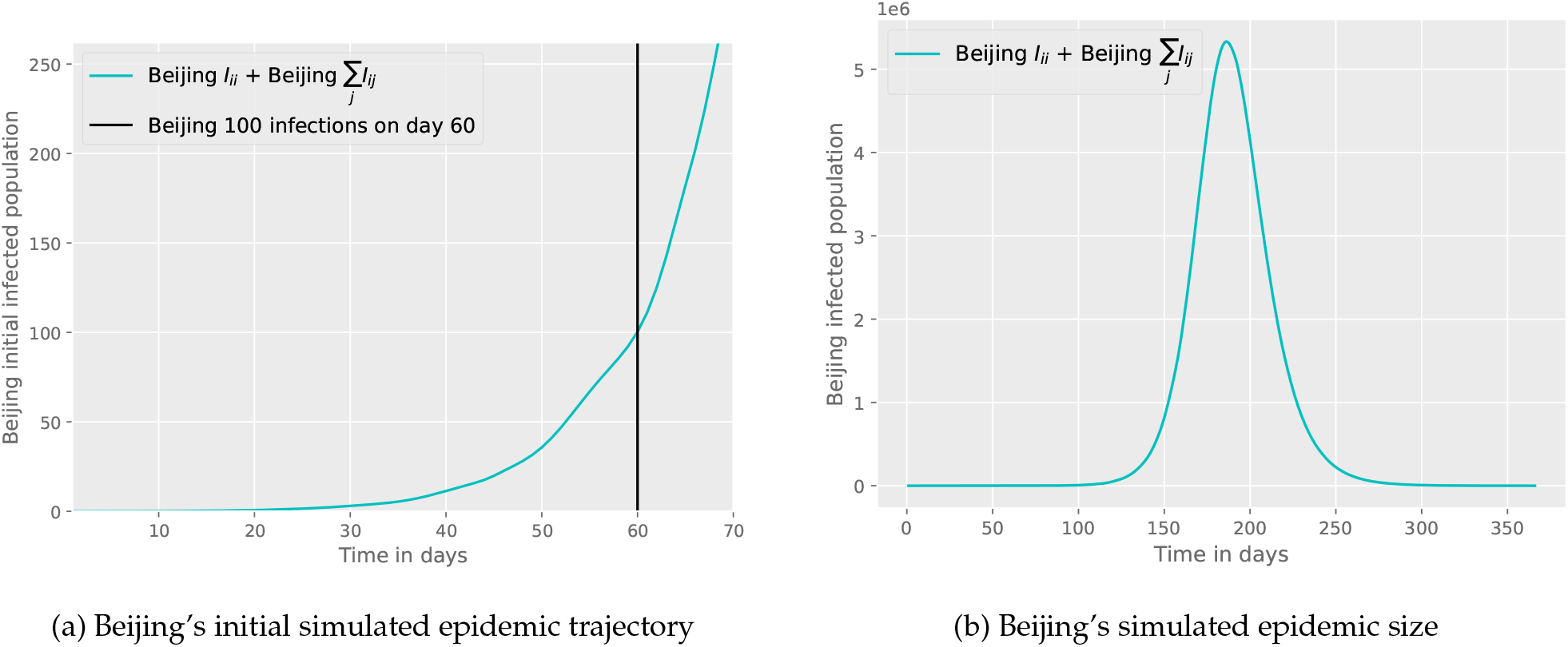
Beijing’s simulated epidemic size. Beijing’s initial epidemic size based on our metapopulation SEIR, taking 60 days to reach 100 local cases.

By employing parameters for a virus resembling SARS-CoV-2 and utilizing real transport flow data, our SEIR metapopulation model demonstrates that the spread occurs swiftly across the population with a significant peak of cases 186 days after the introduction of the disease, with a total of 5.33 million individuals infected in Beijing with no infection control measures (Figure 3b).

We show that in such a highly connected network, travel time from a location is the most important parameter that determines how fast the infection can spread. However, we also noted that in such a highly connected graph that as the travel time increases its impact on the speed of viral spread throughout the metapopulation has a decreasing effect (Figure 4). Our simulations show that Beijing will record 100 infections with an asymptote at approximately 74 days no matter what the travel time from the location to Beijing is once it is above a travel time threshold (Figure 4). These results were largely insensitive to changes in *R*_0_ (see Figure 5).

**Figure 4.**
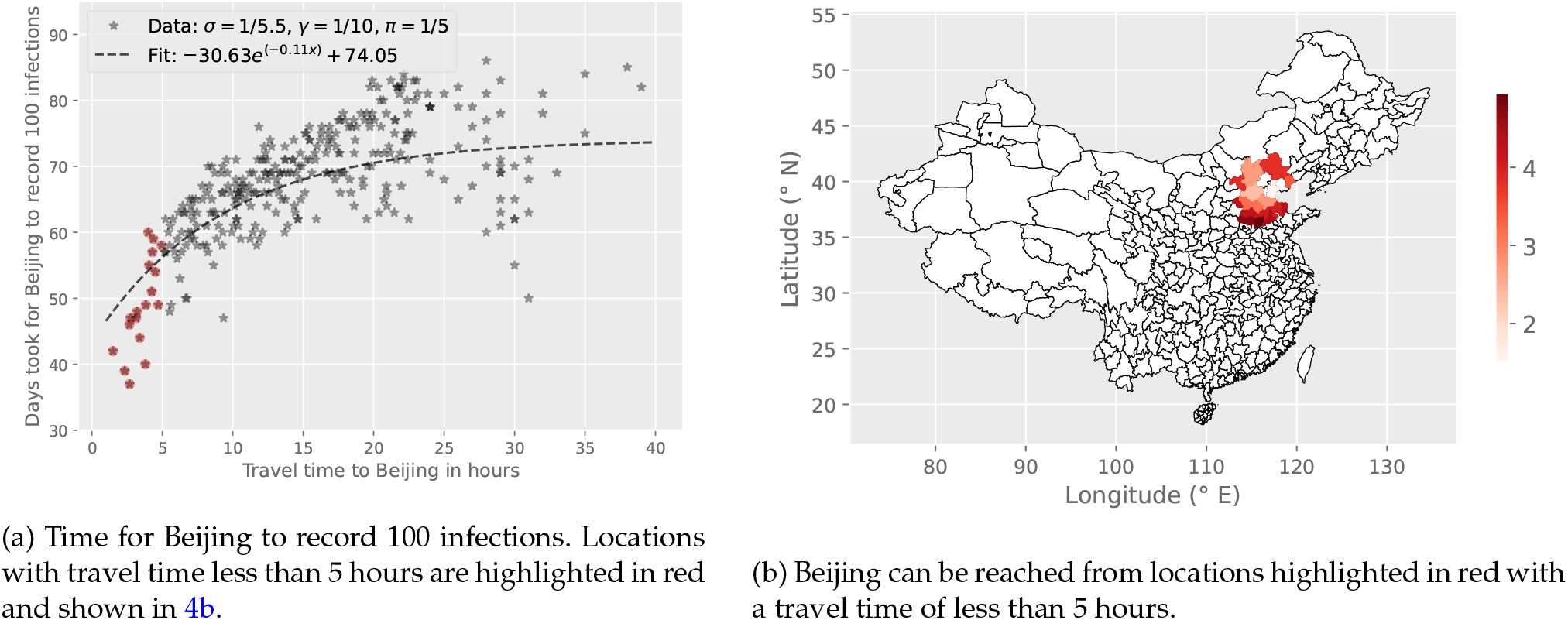
Time for Beijing to record 100 infections following infection introduction in each location in the simulated metapopulation SEIR model. Simulations start with 100 initial infections at each location. Locations within a travel time of less than 5 hours from Beijing deviate from the fit, which may be attributed to the fact that people move more frequently at shorter distances, whereas our assumption is of a constant return rate (*π*) is 1*/*5days.

**Figure 5.**
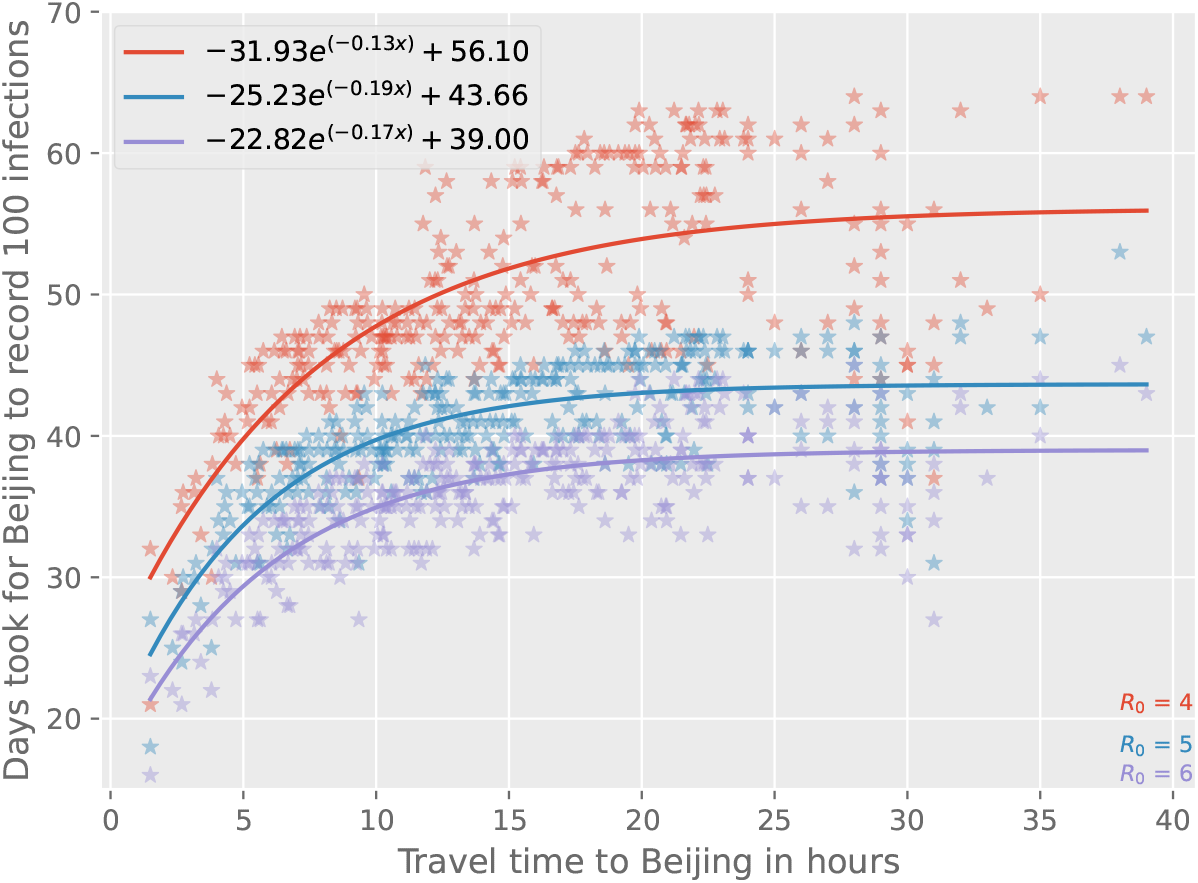
This figure depicts the same data as shown in Figure 4a, but with varying values of *R*_0_.

Principal component analysis (PCA) reveals that six PCs explained 99.45% of the data variation, with PC1 36.7% and PC2 29.35%. The PCs broadly separated travel and distance related factors from ranking statistics and population (PC1) or travel and distance related factors and centrality metrics from weighted rankings (PC2) (Figure 6). Euclidean distance to Beijing, Travel time to Beijing from Google maps, and Travel time to Beijing from Open Street map have positive loadings in PC1, and Distance to Beijing, Travel time to Beijing from Google maps, and Travel time to Beijing from Open Street map are correlated and have similar relationships, whereas Degree Centrality has a negative loading on PC1. Eigenvector Centrality, Betweenness Centrality, and Hubs are another set of non-unique features, so we can select one of them, such as Betweenness Centrality, for further analysis. PageRank is a weak feature in the analysis; however, network ranking statistics such as Weighted Personalized PageRanks and population have positive loadings in PC2. Seven unique metrics identified from the PCA analysis are now utilized in the algorithms presented below and created the feature importance plot (Figure 7).

**Figure 6.**
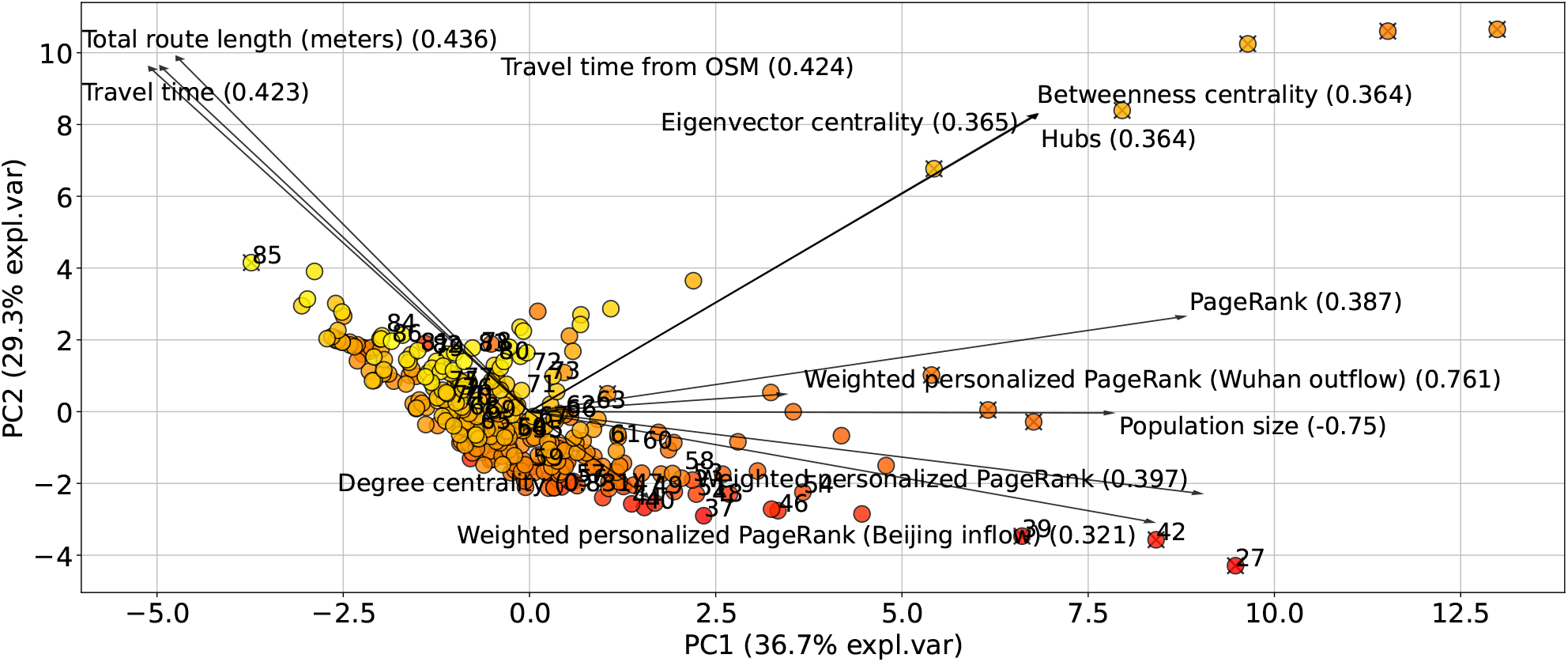
Factors and network metrics that putatively affect the speed of an infection’s spread to Beijing. The Principal Component Analysis shows the relationship between the metrics among the network nodes. From the results of the PCA, we identified unique features, including travel time, degree centrality, betweenness centrality, population size, PageRank, Weighted Personalized PageRank (Wuhan outflow), and Weighted Personalized PageRank (Beijing flow). These unique features were used in machine learning methods to determine their variable importance. See text for details. The numbers in the bracket represent the loading magnitude for each feature.

**Figure 7.**
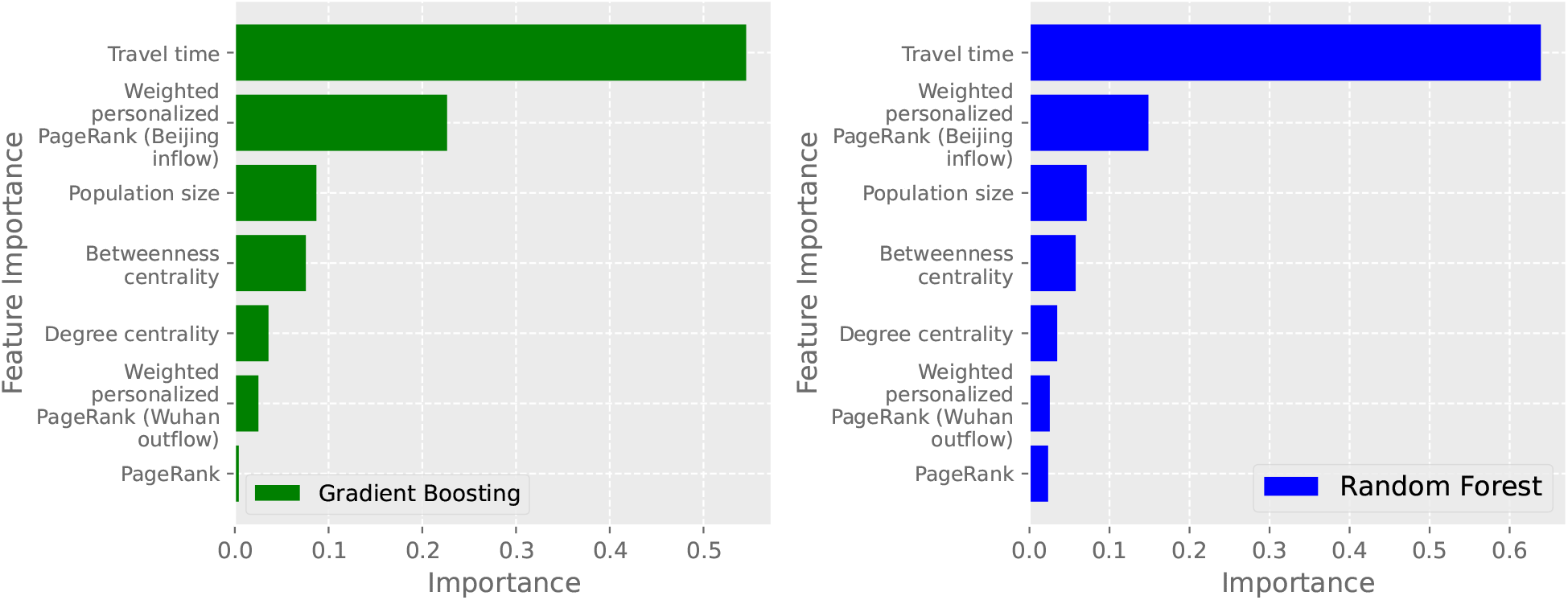
The most important factors affecting the speed of an infection’s spread to Beijing calculated using two different machine learning algorithms, gradient boosting regression trees (green) and random forest (blue).

Both machine learning analyses showed the same rankings. Both showed that travel time was the most important variable, but after travel time Weighted Personalized PageRank for Beijing flow was the most important in determining infection spread to Beijing, with population size next, then betweenness centrality.

Altering the return rate to from 1*/*1 to 1*/*30 showed that after approximately 1*/*12, the return rate has limited impact on the spread of infection throughout the network, as evidenced by the asymptote in Figure 8a bottom panel. Kernel density estimation on the set of points with a derivative of days taken to record 100 infection with respect to 1*/π* greater than *−*1.0, *−*0.5, and *−*0.1 shows changing patterns, especially at *−*1.0. The resulting histograms are shown Figure 8b.

**Figure 8.**
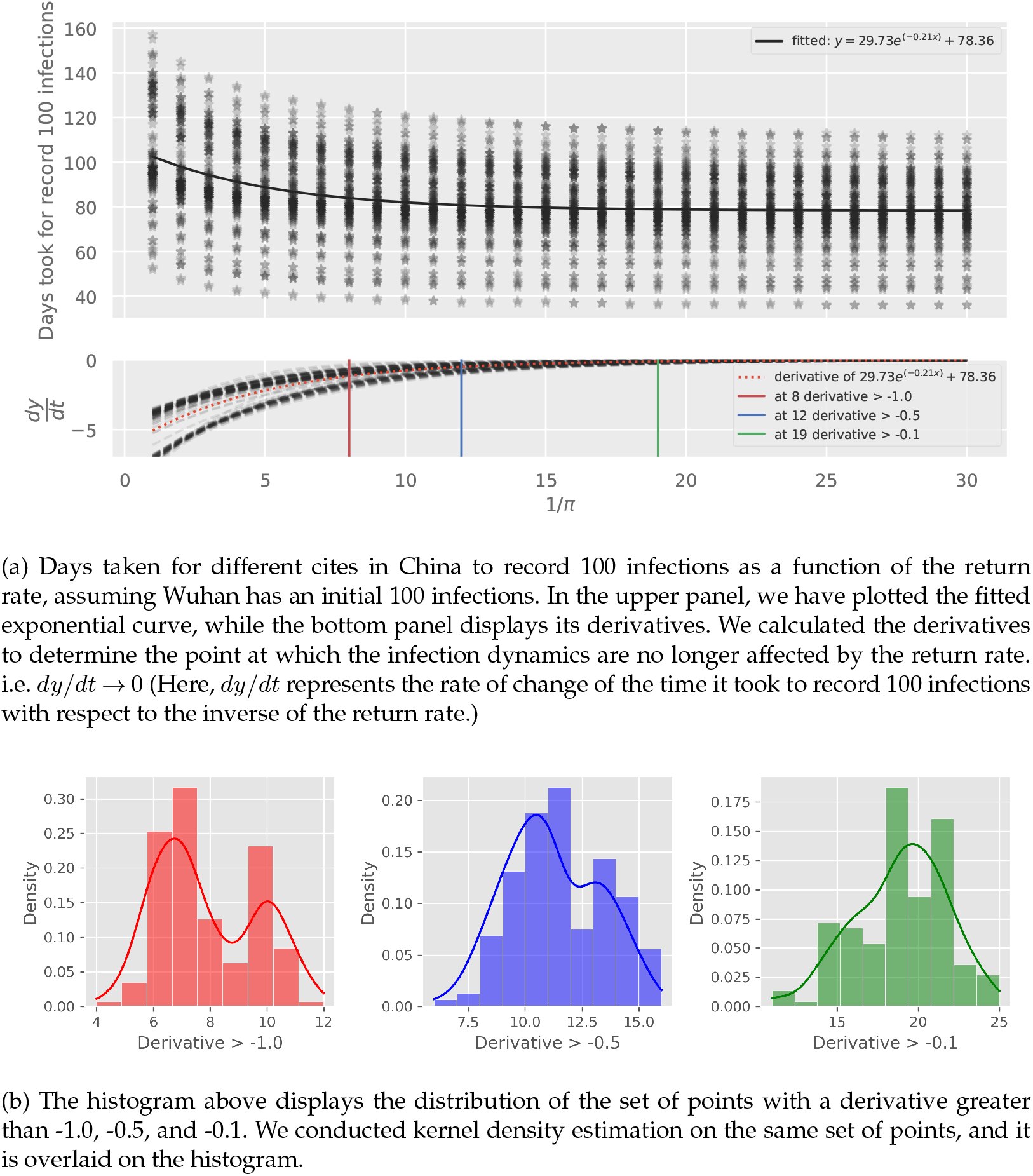
Return rate variation and its effect in infection spread. (a) Variation in return rate and response from simulations for the number of days to reach 100 infections and its asymptote values according to different derivative marks. (b) Kernel density estimation for the fitted values.

In this highly connected network, travel restrictions had to be severe to limit spread. Reducing more than 70% of flow from Wuhan led to just a 10 day increase in the time it took for Beijing to reach 100 cases (from 59 to 69 days) (Table 4). It required an 90% reduction before spread slowed substantially (a 21 day decrease).

**Table 4:**
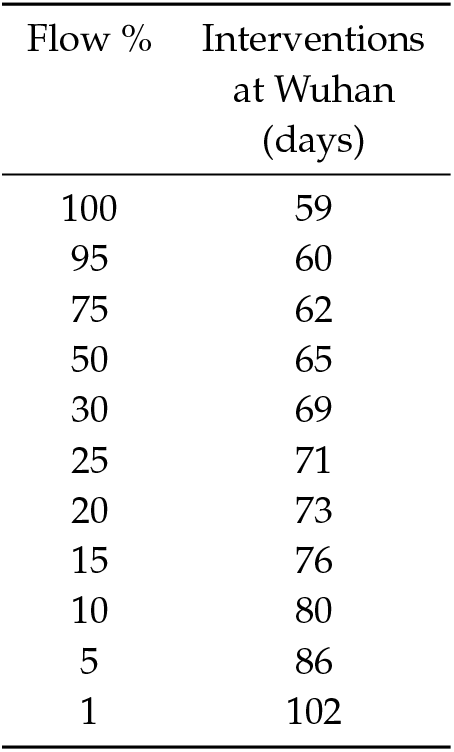
Time in days for Beijing to record 100 infections, when we implement interventions at Wuhan. We changed the outflow from Wuhan from 100% to 1% and ran the simulations to see how much time it would take for Beijing to record 100 infections for both scenarios.

## 4. Discussion

We developed a data-driven metapopulation *SEIR* model to study the transmission dynamics of COVID-19-like diseases in highly connected countries like China and how different network properties impact infection spread. We found the network representing the metapopulation to be densely connected. The spread of SARS-CoV-2-like viruses is rapid throughout such highly connected networks, so much so that an asymptote is reached where travel times stop being important predictors of spread (Figure 4). Further, in such highly connected networks, we found transmission throughout a network when seeded into a location is largely insensitive to non-pharmaceutical interventions, unless human movement was severely restricted (Table 4).

The importance of connectivity for infection spread has previously been noted, including for SARS-CoV-2 [3, 4, 12, 50–52], but here we quantify the importance of specific network properties including data-driven human movement and travel times to infer connectivity. Our analyses are likely globally relevant, as evidenced by the very rapid spread of SARS-CoV-2 leading to the COVID-19 pandemic, once infection had escaped the city of Wuhan, despite China’s strict human movement restrictions [5, 6], which largely continued until recently. Indeed, Wu and colleagues suggested a 50% reduction in inter-city mobility would have a negligible effect on epidemic dynamics [3], and our work supports and extends that, showing only severely limited flow begins to limit spread (Table 4). Chinazzi and colleagues further demonstrated that travel quarantines introduced in Wuhan on 23 January 2020 only delayed the SARS-CoV-2 epidemic progression by 3 to 5 days within China, though it had a greater effect on international spread, presumably because international spread is more managed (e.g. via flights). Few other countries or regions succeeded in getting close to eliminating SARS-CoV-2 using non-pharmaceutical methods, with New Zealand, Australia, Hong Kong, and Singapore among the few [53]. For example, New Zealand, a small, geographically isolated country, greatly reduced domestic infection introductions through both massive reductions in international travel with quarantine and domestic travel restrictions until national vaccination campaigns reached successful outcomes [54]. However, these were only likely feasible (and possibly socially acceptable) with earlier, less transmissible variants of SARS-CoV-2 which have now been largely superseded [55, 56]. Notably, given China’s recent change from a “zero COVID-19” policy, our model predicted that without interventions, China could have 5.4 million infected people after 233 days using early variant (wild-type) epidemiological parameters. More infectious variants that are currently circulating would be likely to cause many more infections than this.

Our machine learning models found travel time to be the most important factor in determining spread, despite its impact plateauing after a point (Figure 8). We also show that once return rates fall within the incubation period of an infection, the transmission dynamics across the network change as commuters can return before infection occurs within the locations travelled to, as shown by the bimodal distributions of the derivative of days took to record 100 infection with respect to 1*/π* in Figure 8b. After travel time, Weighted Personalised Page Rank was most important, with greater importance than population size or Betweenness Centrality, providing evidence that human connectivity is a very strong driver of early infection spread. The regional spread of influenza infection was found to similarly correlate more closely with rates of movement of people to and from workplaces than with geographical distance in the United States, with a similar rapid decay of commuting up to around 100 km and a long tail of rare longer range flow [57].

Together, our work suggests there needs to be multiple approaches to reducing infection transmission for pathogens, because limiting movement alone among highly connected populations is ineffective for highly transmissible infections. Other non-pharmaceutical methods include contact tracing and isolation, which has varying degrees of success depending on the systems and infections (e.g. [58]), and pharmaceutical methods which mostly comprises immunisation, or a combination of these (e.g. [59]). Immunisation, however, is pathogen specific and to date universal vaccines for infections such as influenza [60, 61] and coronaviruses [62] do not exist and novel infectious agents may emerge. Therefore, our work further highlights the need for “primary prevention” of infection emergence at the areas of high risk [63], rather than a “preparedness-response” approach that aims to limit spread in human populations after the emergence of novel infections [64].

Our analyses has several limitations. We used a deterministic model because we are modelling large populations, but stochasticity can be important, particularly for infection establishment and spread among smaller populations, which might be more relevant for understanding the initial phases of infectious disease emergence. Future analyses of early introduction dynamics and of smaller communities simulating the early introduction of infection could be interesting. Similar to the use of deterministic models, we assume homogeneous mixing within populations. However, structural factors such as age can impact transmission dynamics by altering attack rates (the total number of infected individuals) and the basic reproduction number (*R*_0_), which represents the number of cases generated by a typical index case in a fully susceptible population [65]. There are numerous advances in modeling human mobility, however studies increasingly show that patterns are generalisable across scales (i.e., within and between cities and countries) [66–69]. We also do not allow loss of immunity or the emergence of new escape variants, which allows reinfection and alters the transmission dynamics over time, but this is less of a concern for our analysis as we are interested in the initial stages of spread [70]. One additional limitation is that human movement data is only available for the top 100 cities connected to each city in mainland China. Nevertheless, our Baidu data covers 85-99.9% (with median of 92.87%) of movement among cities by using the 100 most connected cities to the 340 cities. Because of that, we believe our analysis provides a comprehensive analysis of the mobility patterns in this hyper-connected network.

## 5. Conclusions

Human movement is fundamental to our way of living and to infectious disease transmission [6, 57, 71]. Our data-driven metapopulation SEIR model of transmission dynamics of COVID-19 like diseases in China shows the spread of SARS-CoV-2-like viruses is rapid throughout such highly connected networks, so much so that an asymptote is reached where travel times stop being important predictors of spread. Our analyses found travel time to be the most important factor in determining spread, despite its impact plateauing after a point, with network metrics weighted by movement of people, modelled here through the Weighted Personalized PageRank, the next most important factor, providing evidence that human connectivity is a driver of infection spread, which can inform mitigation in the future.

## Data Availability

All data produced in the present study are available upon reasonable request to the authors

https://github.com/rejusam/SEIR_metapopulation_China.git

## A. Appendix 1

### Ethics

Ethics approval was not required for this study.

### Data Accessibility

https://github.com/rejusam/SEIR_metapopulation_China.git

### Authors’ Contributions

R.S.J.: conceptualization, investigation, methodology, formal analysis, writing-original draft, Data curation, Software, Validation, Visualization; J.C.M.: conceptualization, formal analysis, investigation, writing—review and editing; R.L.M.: investigation, methodology, validation, writing—review and editing; D.T.S.H.: conceptualization, funding acquisition, investigation, methodology, project administration, writing-original draft.

### Competing Interests

We declare no conflicts.

### Funding

RSJ, RLM, and DTSH were supported by Bryce Carmine and Anne Carmine (née Percival), through the Massey University Foundation (RM22688) and DTSH the Percival Carmine Chair in Epidemiology and Public Health and Royal Society Te Apū rangi RDF-MAU1701. JCM would like to acknowledge the start-up funding La Trobe University, Australia provided.

## Acknowledgements

Massey University’s subscription to New Zealand eScience Infrastructure (NeSi) enabled us to use high-performance computing facilities.

